# Clinical and immunoserological status 12 weeks after infection with COVID-19: prospective observational study

**DOI:** 10.1101/2020.10.06.20206060

**Authors:** Lucía Valiente-De Santis, Inés Pérez-Camacho, Beatriz Sobrino, Gracia Eugenia González, Juan Diego Ruíz-Mesa, Antonio Plata, Ignacio Márquez-Gómez, Marcial Delgado-Fernández, Manuel Castaño, Francisco Oñate, Francisco Orihuela, Begoña Palop, José María Reguera

**Affiliations:** IBIMA. Department of Infectious Diseases; Regional University Hospital of Málaga. Department of Infectious Diseases; Regional University Hospital of Málaga. Department of Pneumology; Regional University Hospital of Málaga. Department of Microbiology

## Abstract

**Objectives:** to undertake a multidisciplinary follow-up at 12 weeks after an acute episode of COVID-19 to assess the functional status, persistence of symptoms and immunoserological situation.

**Methods:** this prospective, observational, single-centre study included outpatients reviewed 12 weeks after an acute infection with SARS-CoV-2. The clinical evaluation included data about the acute episode and epidemiological and clinical variables. The patients were classified as symptomatic or asymptomatic depending on the persistence or otherwise of symptoms. All the patients underwent a full blood test and serology for SARS-CoV-2, as well as imaging tests and spirometry if needed.

**Results:** The mean age of the 108 patients was 55.5 (SD: 15.4) years and 27.8% were health-care workers; 75.9% presented some type of symptoms, with dyspnoea being the most common. A D-dimer >500 ng/mL was detected in 32 (31.4%) patients. All the patients had antibodies against SARS-CoV-2. Being a health-care worker was associated with symptom persistence, with age ≥65 years being a protective factor.

**Conclusions:** The persistence of symptoms in patents with COVID is usual 12 weeks after the acute episode, especially in patients <65 years and health-care workers. All our patients had developed antibodies by 12 weeks.

## Introduction

Respiratory infection (COVID-19) caused by SARS-CoV-2 is characterised by asthenia, fever, dry cough, anosmia, arthromyalgia and dyspnoea, and less commonly by nasal congestion, headache, odynophagia, vomiting and diarrhoea. One week into the course of the disease some patients develop a very aggressive inflammatory response that can lead to respiratory failure secondary to adult respiratory distress, septic shock or coagulation disorders (1,2), which in turn can produce venous thromboembolism (3). Reports also exist of involvement of the heart (4,5), nervous system (6), liver, kidney and skin (7) and eyes (8). Whether these lesions heal definitively or whether they leave persisting damage is unknown. Accordingly, it seems advisable to undertake monitoring and follow-up of patients who have had COVID-19 to determine any possible sequelae (9).

Few data are currently available about the immunoserological response in convalescing patients. Studies have suggested that the serological response with effect from 3 months after symptom onset could reach 100% (10). Nevertheless, little information exists about the duration of the presence of antibodies in COVID-19 patients. We therefore undertook a multidisciplinary follow-up of all COVID-19 patients seen at our hospital to determine their functional and immunoserological status, assess the presence of possible sequelae and evaluate their course.

## Material and Methods

### Patients

This prospective, observational, single-centre study included the first patients seen at the outpatients’ office 12 weeks after acute SARS-CoV-2 infection diagnosed between 14 March and 15 April 2020 and who were discharged after hospital admission or after being seen at the emergency service of the Regional University Hospital in Malaga, Spain but who did not require admission, just home confinement during the acute phase. The patients were classified during the acute phase as a confirmed case (symptoms compatible with COVID-19 and positive result for the SARS-CoV-2 polymerase chain reaction (PCR) (Cobas 6800, Roche) in respiratory samples, or a suspected case (symptoms compatible with COVID-19 and negative PCR) (11).

All the patients were contacted by telephone 12 weeks after the acute phase to undertake a first assessment of their clinical status and make an office appointment.

### Clinical assessment

The clinical assessment was undertaken jointly by an infectious diseases specialist and a pulmonologist. The symptoms during the acute phase were classified as severe if the patient required hospital admission and mild if not. Comorbidity was assessed with the age-adjusted Charlson index. The patients was considered to have received specific treatment for COVID-19 if any of the following drugs were given for at least 24 hours: lopinavir/ritonavir, hydroxychloroquine or chloroquine (with or without azithromycin), tocilizumab, anakinra and steroids. A clinical history was taken of the current symptoms and all the patients underwent a physical examination. The patients were asked about the presence of fever (temperature >37.5ºC), cough, dyspnoea, chest pain, palpitations, arthromyalgia, asthenia, diarrhoea, headache, anosmia, dysgeusia and psychological or cognitive changes (anxiety, mood disorders, insomnia, loss of memory, difficulty concentrating). Patients were considered to be symptomatic when they presented at least one of the symptoms and asymptomatic otherwise. SARS-CoV-2 infection was ruled out in suspected cases with a negative COVID-19 PCR and negative results for IgM and IgG on the serological study at 12 weeks in the absence of immunosuppression. The patients were also classified according to whether they were health-care workers or not.

### Complementary studies

Prior to the office visit all the patients were shown to have a negative SARS-CoV-2 PCR in nasopharyngeal exudate. The following complementary tests were all undertaken at the same time: total leukocytes and lymphocytes, haematocrit, platelets, lymphocyte subsets, D-dimer, creatinine, aspartate aminotransferase, alanine aminotransferase, gamma glutamyl transferase, alkaline phosphatase (AP), lactate dehydrogenase (LDH), ferritin, interleukin 6 (IL-6), immunoglobulins IgA, IgM and IgG and C3 and C4 fractions plus a plain chest radiograph.

Patients considered to require it also underwent respiratory function tests with diffusion testing, computerised tomography (CT) or CT angiography of the chest, echo-Doppler of the legs, ventilation/perfusion gammagraphy and echocardiogram.

### Serological study

The patients also underwent blood tests for IgG and IgM + IgA antibodies against SARS-CoV-2 with the kits COVID-19 VIRCLIA^®^ IgG MONOTEST (Vircell) and COVID-19 VIRCLIA^®^ IgM+IgA MONOTEST (Vircell) based on chemiluminescence (CLIA, ChemiLuminescent ImmunoAssay). The following antigens were used in these kits: the spike protein (S protein) and the nucleocapsid protein (N protein). The use of both types of antigen gives the tests excellent sensitivity, in addition to that of the CLIA technique. For IgM a titre of 0.4-0.6 was considered indeterminate and a titre >0.6 as positive. For IgG a titre of 1.4-1.6 was considered indeterminate and a titre >1.6 as positive. In both cases lower titres were considered negative.

Lymphocyte subset analysis was with the BD Multitest 6-Color TBNK Reagent panel, which includes T lymphocytes (CD4, CD8, CD16 natural killer), B lymphocytes (naive, with and without isotope switching, transitional and plasma) and monocyte subsets (CD14, CD16 effector and regulatory).

### Posterior follow-up

All the cases classified during the acute phase as suspected were reclassified as confirmed if they presented a positive SARS-CoV-2 serology at 12 weeks. All the patients will be given office appointments each three months for a minimum follow-up of one year.

### Statistical study

The continuous variables, expressed as the mean (standard deviation, SD), were compared with the Mann-Whitney U test, and the categorical variables, expressed as number (%), were compared with the χ^2^ or Fisher’s exact tests. A P value <0.05 was considered significant. The statistical analysis was done with SPSS 22.0.0.0.

## Results

Of the original 116 patients contacted 3 (2.5%) refused to attend. Of the remaining 113, 5 (4.4%) were excluded as they proved not to be COVID cases. Table 1 shows the baseline characteristics of the patients during the acute episode. The mean age of the 108 patients was 55.5 (SD: 15.4) years, and 82 (75.9%) had some sort of symptom at the time of office revision.

**Table 1.** Baseline characteristics of the patients during the acute episode

| Characteristic | Total<br>N = 108 | Symptomatic<br>N = 82 | Asymptomatic<br>N = 26 | p |
| --- | --- | --- | --- | --- |
| Sex, N (%) |  |  |  |  |
| · Female | 60 (55.6) | 47 (57.3) | 13 (50) | NS |
| · Male | 48 (44.4) | 35 (42.7) | 13 (50) |  |
| Age ≥65 years | 29 (26.9) | 17 (20.7) | 12 (46.2) | 0.011 |
| Health-care worker, N (%) | 30 (27.8) | 28 (34.1) | 2 (7.7) | 0.009 |
| Charlson > 3, N (%) | 21 (19.4) | 11 (13.4) | 10 (38.5) | 0.005 |
| Acute symptoms, N (%) |  |  |  |  |
| · Mild | 64 (59.3) | 48 (58.5) | 16 (61.5) | NS |
| · Severe | 44 (40.7) | 34 (41.5) | 10 (38.5) |  |
| ICU during acute episode, N (%) | 4 (3.7) | 3 (3.7) | 1 (3.8) | NS |
| Treatment during acute episode | 86 (79.6) | 63 (76.8) | 23 (88.5) | NS |
| Laboratory results during acute episode |  |  |  |  |
| Lymphocytes during acute episode* ** |  |  |  |  |
| · Lymphopenia | 33 (33) | 24 (32.4) | 9 (34.6) | NS |
| · No lymphopenia | 67 (67) | 50 (67.6) | 17 (65.4) |  |
| D-dimer during acute episode* |  |  |  |  |
| · D-dimer >500 | 71 (77.2) | 52 (74.3) | 19 (86.4) | NS |
| · D-dimer <500 | 21 (22.8) | 18 (25.7) | 3 (13.6) |  |
| Ferritin during acute episode* |  |  |  |  |
| · Ferritin ≤1000 ng/L |  |  |  | NS |
| · Ferritin >1000 ng/L | 38 (76) | 29 (74.3) | 9 (81.8) |  |
|  | 12 (24) | 10 (25.6) | 2 (18.2) |  |
| LDH during acute episode* |  |  |  |  |
| · LDH ≤246 U/L | 20 (35.1) | 17 (42.5) | 3 (17.6) | NS |
| · LDH >246 U/L | 37 (64.9) | 23 (57.5) | 14 (82.4) |  |

\*De los patients que estaba disponible
\*\*Cifra de linfocitos menor de 900
ICU, intensive care unit; LDH, lactate dehydrogenase

Table 2 shows the symptoms, with a predominance of respiratory symptoms. Concerning the number of symptoms, 18.6% reported just one, 19.5% two and 39.8% three or more symptoms.

**Table 2:** Symptoms 12 weeks after the acute episode

|  |  |
| --- | --- |
| Symptoms | N (%) |
|  | 82 (75.9) |
| Dyspnoea | 60 (55.6) |
| Asthenia | 48 (44.9) |
| Cough | 28 (25.9) |
| Chest pain | 28 (25.9) |
| Palpitations | 24 (22.2) |
| Headache | 10 (9.3) |
| Anosmia | 10 (9.3) |
| Dysgeusia | 5 (5.6) |
| Fever | 4 (3.7) |
| Chills | 4 (3.7) |
| Arthromyalgia | 3 (2.8) |
| Hair loss | 3 (2.8) |
| Diarrhoea | 2 (1.9) |
| Psychological and cognitive disorders: | 18 (16.7) |
| · Anxiety | 7 (6.4) |
| · Sadness | 7 (6.4) |
| · Insomnia | 2 (1.9) |
| · Loss of memory | 2 (1.9) |
| · Difficulty concentrating | 2 (1.9) |

A blood test was performed in 102 (94.4%) of the 108 COVID-19 patients. Table 3 shows the main changes, highlighting an increase in D-dimer >500 ng/mL in 32 (31.4%) patients. No significant differences were seen according to the presence or absence of symptoms

**Table 3:**
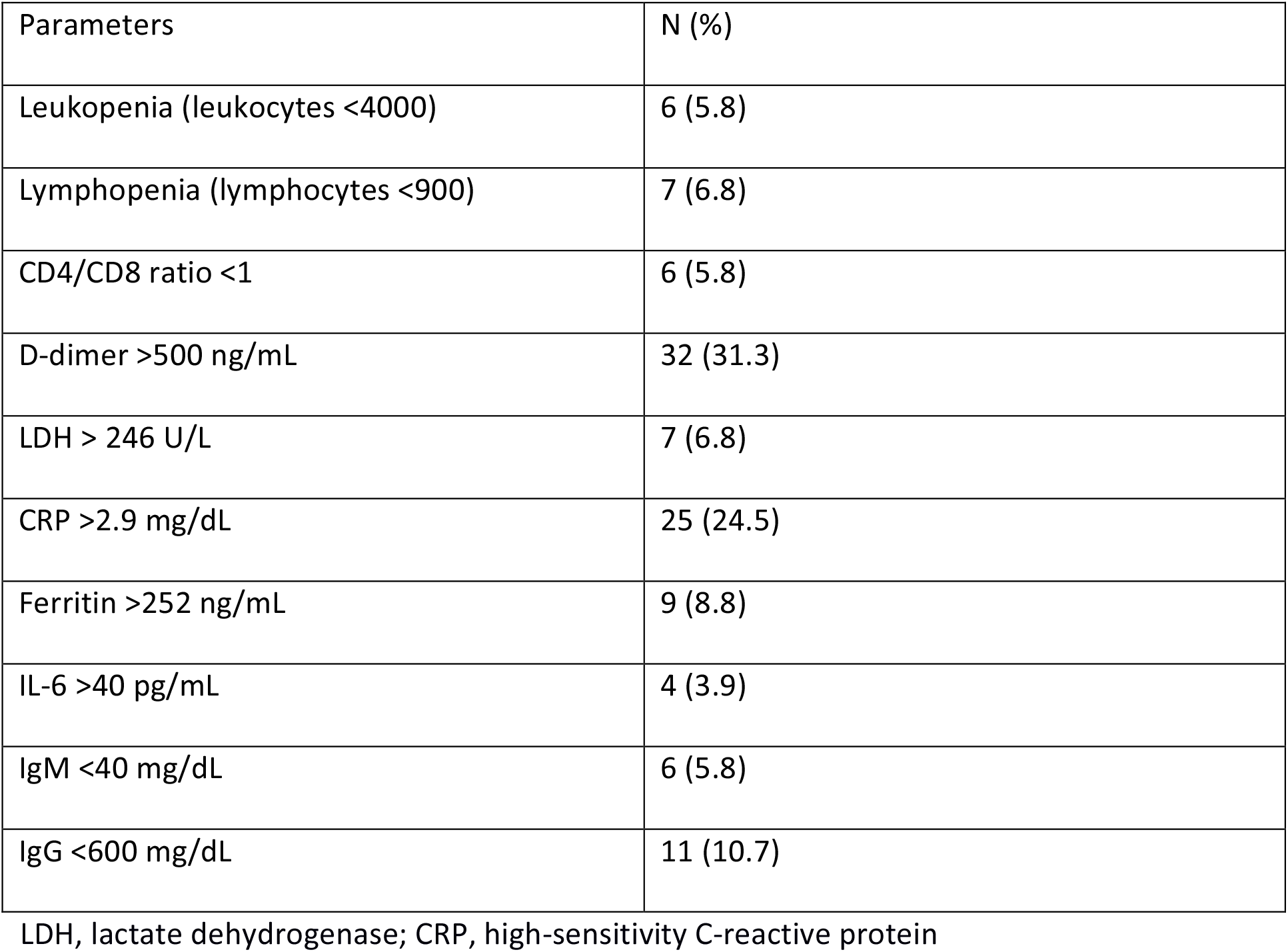
Main results of the laboratory studies

| Parameters | N (%) |
| --- | --- |
| Leukopenia (leukocytes <4000) | 6 (5.8) |
| Lymphopenia (lymphocytes <900) | 7 (6.8) |
| CD4/CD8 ratio <1 | 6 (5.8) |
| D-dimer >500 ng/mL | 32 (31.3) |
| LDH > 246 U/L | 7 (6.8) |
| CRP >2.9 mg/dL | 25 (24.5) |
| Ferritin >252 ng/mL | 9 (8.8) |
| IL-6 >40 pg/mL | 4 (3.9) |
| IgM <40 mg/dL | 6 (5.8) |
| IgG <600 mg/dL | 11 (10.7) |

LDH, lactate dehydrogenase; CRP, high-sensitivity C-reactive protein

Concerning imaging tests, 89 (82.4%) patients had a control chest radiograph at 12 weeks after the acute episode. In 56 (62.9%) this was normal, in 24 (26.9%) the evolution of the images was favourable and in 9 (10.1%) the radiological alterations seen during the acute phase had persisted or worsened. A chest CT was done in 37 (41.5%) patients; it was normal in 7 (18.9%) and pathological in the rest: 24 (64.9%) had ground-glass opacity, 3 (8.1%) a pattern of pulmonary fibrosis, 2 (5.4%) pulmonary thromboembolism and 1 (2.7%) residual lobar infiltrate.

Spirometry was performed in 32 (29.6%) patients. In 23 (71.9%) the result was normal, 4 (12.5%) had an obstructive pattern, 3 (9.4%) a mixed pattern and in 2 (6.3%) the alterations were interpreted as secondary to poor collaboration. None of the baseline characteristics was associated with radiological or respiratory function changes.

The serological test was not undertaken in 3 (2.7%) of the 108 patients. Table 3 shows the serological response of the remaining 105 COVID-19 patients. All patients had a serological response (IgG in 98.1%) and at 12 weeks 55% were positive for both IgM and IgG antibodies (Table 4).

**Table 4:** Serological response

| Antibodies, N (%) | Total | Symptomatic | Asymptomatic | p |
| --- | --- | --- | --- | --- |
| IgM positive | 60 (57.1) | 45 (56.3) | 15 (60) | NS |
| IgM negative | 35 (33.3) | 28 (35.5) | 7 (28) | NS |
| IgM indeterminate | 10 (9.5) | 7 (8.8) | 3 (12) | NS |
| IgG positive | 103 (98.1) | 79 (98.8) | 24 (96) | NS |
| IgG negative | 2 (9.1) | 1 (1.3) | 1 (4) | NS |
| IgM and IgG positive | 58 (55.5) | 44 (55) | 14 (56) | NS |

Being a health-care worker was associated with symptom persistence (OR = 4.79 [95% CI = 1.02-22.38], p=0.046), with age ≥65 years being a protective factor (OR = 0.33 [95% CI = 0.12-0.87], p=0.026) (Table 5).

**Table 5:** Risk factors for persistence of symptoms

| Variable | OR multivariate analysis (95% CI) | p |
| --- | --- | --- |
| Age >65 years | 0.33 (0.12-0.87) | 0.026 |
| Health-care worker | 4.79 (1.02-22.38) | 0.046 |
| Mild or severe acute episode | - | 0.087 |
| Charlson > 3 | - | 0.130 |
| D-dimer >500 ng/mL | - | 0.317 |
| Specific treatment for COVID-19 | - | 0.435 |

## Discussion

Few studies examining the course of patients with COVID-19 have included data about possible sequelae and the immunoserological response against SARS-CoV-2. In our study most of the infected patients (75.9%) in our preliminary SARS-CoV-2 cohort continued to have symptoms 12 weeks after the acute episode. This coincides with the results reported by Carfi et al., who assessed the presence of residual symptoms in 143 patients after a mean of 60 days following the acute episode (12); 87.4% of the patients reported persistence of symptoms.

Dyspnoea was the predominant symptom in most of our patients. A retrospective study comprising 50 patients assessed one month after the acute episode found that over half presented worsened lung function, mostly consisting of restrictive and diffusion alterations, generally mild (13). Likewise, the persistence of diffusion alterations seems more common in patients who have severe pneumonia during the acute episode (14). In our series, the prevalence of alterations detected on spirometry was lower than in the previous series, though our series included more patients with a mild acute episode than the others. Additionally, as spirometry in our cohort was determined by the persistence of symptoms, a lower percentage of patients underwent the test.

Although most of the patients with radiological alterations during the acute phase evolved favourably, 37% still had some degree of persistence of symptoms. The chest CT in these patients confirmed the presence of radiological lesions, most frequently persistence of the ground-glass opacity. This is in contrast to the findings of Zhao et al, who assessed 55 patients three months after acute infection with SARS-CoV-2 and found that 70% has persistence of radiological alterations. However, whereas in our series it was persistence of ground-glass opacity and *crazy paving* in their series it was interstitial thickening compatible with pulmonary fibrosis (15). The three patients with fibrosis in our series had presented severe clinical symptoms, with one having bilateral pneumonia and requiring admission to the intensive care unit, which agrees with the literature (16). Of note is the high number of patients who reported dyspnoea, sometimes with cough and chest pain, but who had no demonstrable respiratory function or radiological alterations. The association of these systemic symptoms together with asthenia and psychological disorders could constitute what has been called post-viral or post-covid chronic fatigue syndrome (17). The presence of anxiety and depression have been reported with SARS (18) and they may persist beyond the duration of the bout. In our series, 16.7% of the patients experienced psychological or cognitive alterations after the acute episode of COVID-19, which is in concordance with the impact that other outbreaks have had for the mental health of the patients and the community in general (19). Nine of our patients required psychiatric assessment.

The most frequent symptoms after respiratory symptoms were neurological symptoms, occurring in about one in every 10 patients, mainly persistence of headache, anosmia and dysgeusia. The presence of these symptoms during the acute infection is common (6,20) and although most cases appear to resolve in less than 4 weeks (21), our results suggest that these alterations may persist in some patients for at least three months.

During the acute phase of SARS-CoV-2 there can be involvement of the myocardium, producing myocarditis. Whilst the prognosis for this appears favourable, we are unaware of the long-term consequences of this myocardial damage, and it may be part of the genesis of the palpitations and chest pain sometimes reported by patients at revision (22). Another important cardiovascular aspect we have noted is the persistence in the rise in D-dimer in over 30% of the patients three months after the acute phase; one of our patients even showed pulmonary thromboembolism on the control CT. This all suggests the need to maintain the patients anticoagulated for an indeterminate period (23). Unfortunately, we are unable to provide a response to this as our study was not designed with this in mind.

Almost all our patients had a humoral immune response by three months after the acute episode. Furthermore, this serological response was independent of the severity of the acute symptoms and the symptoms at the time of revision. Between 2 and 6 weeks after diagnosis of COVID-19 most patients develop antibodies against SARS-CoV-2, as shown in the study by Lin et al. (24) that included 334 patients with COVID-19 confirmed by SARS-CoV-2 PCR. Specific IgM or IgG antibodies were detected in all the patients. It has been suggested that asymptomatic patients could seroconvert to a lesser extent than patients with severe symptoms and could even become negative during the convalescence (25). However, we failed to see this as all our patients had detectable antibodies 3 months after the acute episode. Subsequent follow-up of our patients will show the evolution of the immunoserological response.

Care of patients with COVID-19 has an important impact on the health of health-care workers (26,27). This impact has both physical and mental consequences (27). A retrospective cohort study found that symptoms of infection by COVID-19 in health-care workers are mild or moderate in 91% of cases (28). Nonetheless, our data show that health-care workers more often have residual symptoms that persist after the acute episode. It would, therefore, be recommendable to undertake an evolutionary follow-up of this group after SARS-CoV-2 infection. Moreover, health-care workers have a greater risk of post-traumatic stress, with the younger workers more often developing this condition (26). In our study the patients <65 years had a greater risk of residual symptoms.

In conclusion, 12 weeks after the acute episode residual symptoms are common in patients with COVID, the most frequent being respiratory involvement. Health-care workers and patients <65 years have a greater risk of presenting persistent symptoms after the acute episode. An increase in D-dimer suggests the possibility of a state of hypercoagulabilty persisting after the acute phase. All our patients had developed IgM or IgG antibodies against SARS-CoV-2 by 12 weeks.

## Data Availability

Regional University Hospital of Malaga

## Notes

### Competing Interest Statement

The authors have declared no competing interest.

### Funding Statement

No external funding was received

### Author Declarations

The study was approved by Comite de Etica de la Investigacion Provincial de Malaga

